# Spatial-temporal relationship between population mobility and COVID-19 outbreaks in South Carolina: A time series forecasting analysis

**DOI:** 10.1101/2021.01.02.21249119

**Authors:** Chengbo Zeng, Jiajia Zhang, Zhenlong Li, Xiaowen Sun, Bankole Olatosi, Sharon Weissman, Xiaoming Li

## Abstract

**Background:** Population mobility is closely associated with coronavirus 2019 (COVID-19) transmission, and it could be used as a proximal indicator to predict future outbreaks, which could inform proactive non-pharmaceutical interventions for disease control. South Carolina (SC) is one of the states which reopened early and then suffered from a sharp increase of COVID-19.

**Objective:** To examine the spatial-temporal relationship between population mobility and COVID-19 outbreaks and use population mobility to predict daily new cases at both state- and county- levels in SC.

**Methods:** This longitudinal study used disease surveillance data and Twitter-based population mobility data from March 6 to November 11, 2020 in SC and its top five counties with the largest number of cumulative confirmed cases. Daily new case was calculated by subtracting the cumulative confirmed cases of previous day from the total cases. Population mobility was assessed using the number of users with travel distance larger than 0.5 mile which was calculated based on their geotagged twitters. Poisson count time series model was employed to carry out the research goals.

**Results:** Population mobility was positively associated with state-level daily COVID-19 incidence and those of the top five counties (i.e., Charleston, Greenville, Horry, Spartanburg, Richland). At the state-level, final model with time window within the last 7-day had the smallest prediction error, and the prediction accuracy was as high as 98.7%, 90.9%, and 81.6% for the next 3-, 7-, 14- days, respectively. Among Charleston, Greenville, Horry, Spartanburg, and Richland counties, the best predictive models were established based on their observations in the last 9-, 14-, 28-, 20-, and 9- days, respectively. The 14-day prediction accuracy ranged from 60.3% to 74.5%.

**Conclusions:** Population mobility was positively associated with COVID-19 incidences at both state- and county- levels in SC. Using Twitter-based mobility data could provide acceptable prediction for COVID-19 daily new cases. Population mobility measured via social media platform could inform proactive measures and resource relocations to curb disease outbreaks and their negative influences.

## Introduction

Since the first confirmed case of Coronavirus Disease 2019 (COVID-19) in the United States (US) on January 21, 2020, the countrywide COVID-19 outbreaks have surged quickly. As of December 29, there were 19,566,140 cumulative confirmed cases and 338,769 COVID-19 related deaths in US [1]. South Carolina (SC), a state located in Southeastern US, had the first confirmed cases on March 6, 2020. From March to May, the trends of daily new cases were flat with an average of daily increased cases less than 500. However, after the early implementation of reopening policies, the daily new cases in SC have risen sharply since June. On July 14, the COVID-19 cases in SC surpassed 60,000, with more than 2,200 daily new cases, the second highest increase in one day in the US [2]. Between August and October, the transmission rate slowed down with the further implementation of non-pharmaceutical interventions (NPIs), such as dine-in service restriction and face-covering requirement, but increased steadily after October. By December 29, there were 300,602 reported cases and 5,198 deaths in SC [3]. Given the rapid transmission of COVID-19 in SC, more research is needed to identify potential early predictors of increasing transmission rates which could then be used to inform proactive NPIs to suppress statewide disease transmission.

Population mobility is a potential early indicator of COVID transmission as population mobility reflects the influences (both positive and negative) of NPIs, reopening actions, social distancing practices and public holidays [4-6]. For instance, at the early stage of COVID-19 epidemic, the SC Governor issued a series of NPIs, such as shelter-in-place and school and non-essential business closure, to reduce social interaction. These NPIs showed positive effects in suppressing the statewide COVID-19 spread. Later in May, the reopening policies and public holidays diluted the implementation of NPIs leading to the increased social interactions and statewide COVID-19 spread [7,8]. At present, it may be difficult to measure the real-time impact of reopening policies, public holidays and fidelity of NPIs implementation. Therefore, population mobility could be a proximal indicator allowing for real-time COVID-19 transmission forecasting.

Social media platforms, such as Twitter, collect geospatial information and closely monitor the change of population mobility [9,10]. Indeed, the tremendous volume of user-generated geoinformation from social media helps promote the real-time or near real-time surveillance of population mobility and provides timely data on how population mobility responded to different phases of COVID-19 outbreak, policy reactions, and public holidays [11- 13]. Several studies have leveraged mobility data from social media (e.g., Google, Facebook, Twitter) to investigate the relationships between population mobility and COVID-19 transmission [6,8,14-16]. These studies identified a consistently positive relationship between population mobility and COVID-19 incidence. However, few studies used population mobility as a predictor to forecast further outbreaks and to evaluate the prediction accuracy in addition to correlation analysis. One study by Wang and Yamamoto predicted COVID-19 daily new cases in Arizona using disease surveillance data, Google Community Mobility report, and partial differential equation. They found acceptable prediction for the next 3-day [16]. This study only classified Arizona into three regions (i.e., central, northern, southern of Arizona) and evaluated the prediction accuracy for the next 3-day which did not cover the duration of viral incubation (i.e., 14-day). More studies are needed to investigate the relationship between population mobility using social media data and COVID-19 transmission at both state- and county- levels and over longer timeframes.

Leveraging disease surveillance data and Twitter-based population mobility, the current study aimed to construct time series models of COVID-19 daily new cases, investigate the relationship between them, and evaluate the prediction accuracy of daily new cases for the next two-week window at both state- and county- levels in SC.

## Methods

### COVID-19 incidence data

Cumulative confirmed cases of COVID-19 through November 11, 2020 at both state- and county-levels in SC were collected from The New York Times, which was deposited in Github [17]. Within the study period (March 6, 2020 [date of 1^st^ COVID diagnosis in SC] to November 11, 2020 [251^st^ day]), daily new cases were calculated by subtracting the cumulative confirmed cases of previous day from the total cases for the entire state and its five counties with largest numbers of cumulative confirmed cases (i.e., Charleston, Greenville, Horry, Spartanburg, and Richland). The study protocol was approved by the Institutional Review Boards at the University of South Carolina.

### Population mobility

Population mobility was assessed using the number of people (Twitter users) with moving distance larger than 0.5 mile per day in SC and the selected counties. The methodology of extracting daily population movement (origin-destination flows) from geotagged tweets is discussed elsewhere [18,19]. Briefly, geotagged tweets during the study periods were collected and used for calculation. Only users who post at least twice a day or posted tweets on at least two consecutive days were included in the calculation. Daily travel distance was calculated for each user based on the derived origin-destination flows and used to generate a variable of how many people moved each day (with travel distance larger than 0.5 mile). This method of capturing population mobility through Twitter has been previously validated [18].

### Statistical analysis

First, daily new cases of COVID-19 and population mobility at both state- and county-levels were described using line charts in R version 3.6.3 (The R Foundation, “ggplot” package). Daily new cases and mobility were also described using fives quantiles (i.e., minimum, 25^th^ percentile, 50^th^ percentile, 75^th^ percentile, and maximum) by each month.

Second, Poisson count time series model was used to model the impact of population mobility on the daily new cases of COVID-19 at state-level. Time series models were built at the various time windows. At the first round selection, a total of 17 time windows (by a 7-day increment) were considered including 1 to 7 days, 1 to 14 days,…, and 1 to 119 days. The daily new cases from the 1^st^ to 234^th^ days were used as the training dataset and those from the next 3- day (235^th^ ∼ 237^th^) were used as testing dataset for the purpose of model evaluation. With the smallest prediction error (Formula 1) and good interpretation, the predictive model with the best time window was selected. After the best time window in the first round selection was determined, second and third round selections were conducted to narrow down the time window and obtain the final model with the smallest prediction error. The final model was used to predict the COVID-19 daily new cases for the next 3-, 7-, and 14- days (238^th^ ∼ 251^st^ days). Cumulative difference (Formula 2) between observed and predicted cases and mean absolute percentage accuracy (Formula 3) by each timeframe were reported.^16^

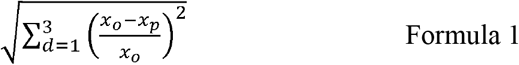

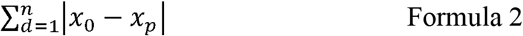

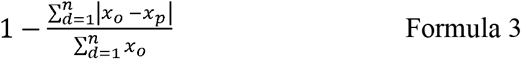

***Notes***: *d*: day; *n*: next 3-, 7-, or 14- days; *o*: observed value; *p*: predicted value; *x*: daily new cases.

Finally, a similar analytic procedure was performed to construct the final model at the county-level for each of the top five counties (i.e., Charleston, Greenville, Horry, Spartanburg, and Richland) in SC. Poisson count time series model was conduct using the R package (“tscount”).

## Results

### Descriptive statistics

Table 1 shows the descriptive statistics of COVID-19 new cases, and Figure 1 shows the changes of COVID-19 daily new cases at both state- and county-levels. By October 31, there were 176,612 cumulative COVID-19 confirmed cases in SC. The cumulative confirmed cases in Charleston, Greenville, Horry, Spartanburg, and Richland were 17,384, 18,021, 12,591, 9,290, and 17,531, respectively. At the state-level, the daily new cases from March to the end of May were less than 500. From June to the middle of July, the daily new cases elevated, with 2,217 new COVID-19 patients on July 14. After that, the transmission rate decreased, with most of the daily new cases less than 1,500. However, since October, the daily new cases steadily increased.

**Table 1.** Descriptive statistics of population mobility and COVID-19 new cases at both state- and county- levels

|  |  | Minimum | 25<sup>th</sup> percentile | 50<sup>th</sup> percentile | 75<sup>th</sup> percentile | Maximum |
| --- | --- | --- | --- | --- | --- | --- |
| **Population mobility** |  |  |  |  |  |  |
| **State-level** |  |  |  |  |  |  |
|  | March | 658 | 809 | 1,010 | 1,109 | 1,438 |
|  | April | 554 | 617 | 670 | 697 | 786 |
|  | May | 630 | 754 | 812 | 848 | 940 |
|  | June | 756 | 848 | 871 | 910 | 993 |
|  | July | 818 | 870 | 896 | 937 | 1,039 |
|  | August | 767 | 828 | 863 | 884 | 1,035 |
|  | September | 784 | 831 | 875 | 907 | 1,021 |
|  | October | 789 | 843 | 898 | 965 | 1,085 |
| **County-level** |  |  |  |  |  |  |
| **Charleston** |  |  |  |  |  |  |
|  | March | 81 | 104 | 126 | 142 | 195 |
|  | April | 62 | 75 | 83 | 92 | 98 |
|  | May | 73 | 93 | 104 | 121 | 140 |
|  | June | 96 | 109 | 116 | 126 | 154 |
|  | July | 95 | 109 | 117 | 121 | 133 |
|  | August | 88 | 99 | 109 | 120 | 134 |
|  | September | 94 | 110 | 116 | 126 | 150 |
|  | October | 95 | 113 | 122 | 132 | 142 |
| **Greenville** |  |  |  |  |  |  |
|  | March | 103 | 115 | 139 | 156 | 177 |
|  | April | 82 | 93 | 106 | 114 | 134 |
|  | May | 100 | 113 | 119 | 127 | 132 |
|  | June | 104 | 117 | 124 | 133 | 162 |
|  | July | 107 | 124 | 135 | 140 | 153 |
|  | August | 111 | 129 | 140 | 146 | 168 |
|  | September | 114 | 128 | 140 | 144 | 158 |
|  | October | 104 | 133 | 138 | 149 | 169 |
| **Horry** |  |  |  |  |  |  |
|  | March | 77 | 84 | 87 | 116 | 158 |
|  | April | 53 | 64 | 71 | 80 | 97 |
|  | May | 76 | 87 | 100 | 128 | 151 |
|  | June | 103 | 113 | 125 | 133 | 162 |
|  | July | 100 | 116 | 123 | 140 | 171 |
|  | August | 89 | 112 | 118 | 137 | 160 |
|  | September | 79 | 96 | 107 | 117 | 143 |
|  | October | 71 | 99 | 105 | 116 | 151 |
| **Spartanburg** |  |  |  |  |  |  |
|  | March | 40 | 67 | 82 | 89 | 106 |
|  | April | 34 | 43 | 47 | 50 | 61 |
|  | May | 47 | 51 | 56 | 62 | 72 |
|  | June | 50 | 62 | 65 | 72 | 78 |
|  | July | 51 | 67 | 76 | 85 | 101 |
|  | August | 50 | 65 | 70 | 77 | 94 |
|  | September | 55 | 62 | 65 | 70 | 74 |
|  | October | 52 | 59 | 67 | 79 | 92 |
| **Richland** |  |  |  |  |  |  |
|  | March | 58 | 76 | 82 | 93 | 120 |
|  | April | 53 | 68 | 73 | 78 | 84 |
|  | May | 61 | 69 | 77 | 84 | 115 |
|  | June | 65 | 77 | 86 | 93 | 105 |
|  | July | 59 | 76 | 82 | 95 | 105 |
|  | August | 72 | 79 | 89 | 95 | 109 |
|  | September | 72 | 82 | 89 | 97 | 125 |
|  | October | 72 | 84 | 92 | 100 | 119 |
| **COVID-19 new cases** |  |  |  |  |  |  |
| **State-level** |  |  |  |  |  |  |
|  | March | 0 | 3 | 18 | 74 | 158 |
|  | April | 62 | 131 | 154 | 204 | 275 |
|  | May | 82 | 129 | 164 | 228 | 467 |
|  | June | 236 | 476 | 757 | 1,115 | 1,755 |
|  | July | 972 | 1,520 | 1,726 | 1,855 | 2,374 |
|  | August | 456 | 722 | 937 | 1,214 | 1,583 |
|  | September | 301 | 624 | 863 | 1,190 | 2,665 |
|  | October | 381 | 789 | 912 | 1,057 | 1,706 |
| **County-level** |  |  |  |  |  |  |
| **Charleston** |  |  |  |  |  |  |
|  | March | 0 | 0 | 1 | 8 | 32 |
|  | April | 0 | 3 | 5 | 12 | 48 |
|  | May | 0 | 1 | 6 | 8 | 23 |
|  | June | 11 | 34 | 69 | 200 | 373 |
|  | July | 85 | 164 | 221 | 303 | 418 |
|  | August | 25 | 53 | 95 | 105 | 218 |
|  | September | 0 | 35 | 46 | 65 | 425 |
|  | October | 13 | 34 | 50 | 61 | 89 |
| **Greenville** |  |  |  |  |  |  |
|  | March | 0 | 1 | 5 | 11 | 18 |
|  | April | 0 | 9 | 19 | 28 | 54 |
|  | May | 7 | 14 | 21 | 33 | 150 |
|  | June | 47 | 71 | 115 | 147 | 245 |
|  | July | 49 | 129 | 167 | 196 | 276 |
|  | August | 14 | 40 | 53 | 95 | 184 |
|  | September | 6 | 41 | 75 | 113 | 289 |
|  | October | 27 | 87 | 107 | 140 | 197 |
| **Horry** |  |  |  |  |  |  |
|  | March | 0 | 1 | 2 | 3 | 5 |
|  | April | 0 | 2 | 5 | 9 | 18 |
|  | May | 0 | 4 | 5 | 10 | 26 |
|  | June | 17 | 47 | 99 | 133 | 221 |
|  | July | 63 | 103 | 145 | 189 | 358 |
|  | August | 16 | 30 | 41 | 56 | 115 |
|  | September | 4 | 20 | 30 | 46 | 70 |
|  | October | 15 | 48 | 73 | 90 | 139 |
| **Spartanburg** |  |  |  |  |  |  |
|  | March | 0 | 0 | 0 | 2 | 7 |
|  | April | 1 | 4 | 6 | 11 | 32 |
|  | May | 1 | 4 | 7 | 14 | 61 |
|  | June | 5 | 18 | 34 | 44 | 72 |
|  | July | 18 | 48 | 63 | 84 | 125 |
|  | August | 11 | 25 | 44 | 62 | 92 |
|  | September | 2 | 18 | 50 | 99 | 215 |
|  | October | 0 | 46 | 78 | 96 | 147 |
| **Richland** |  |  |  |  |  |  |
|  | March | 1 | 3 | 6 | 14 | 37 |
|  | April | 3 | 15 | 25 | 32 | 56 |
|  | May | 5 | 15 | 19 | 26 | 33 |
|  | June | 12 | 44 | 67 | 81 | 155 |
|  | July | 57 | 108 | 138 | 165 | 234 |
|  | August | 39 | 79 | 93 | 124 | 408 |
|  | September | 34 | 77 | 96 | 142 | 766 |
|  | October | 24 | 51 | 67 | 78 | 130 |

**Figure 1.**
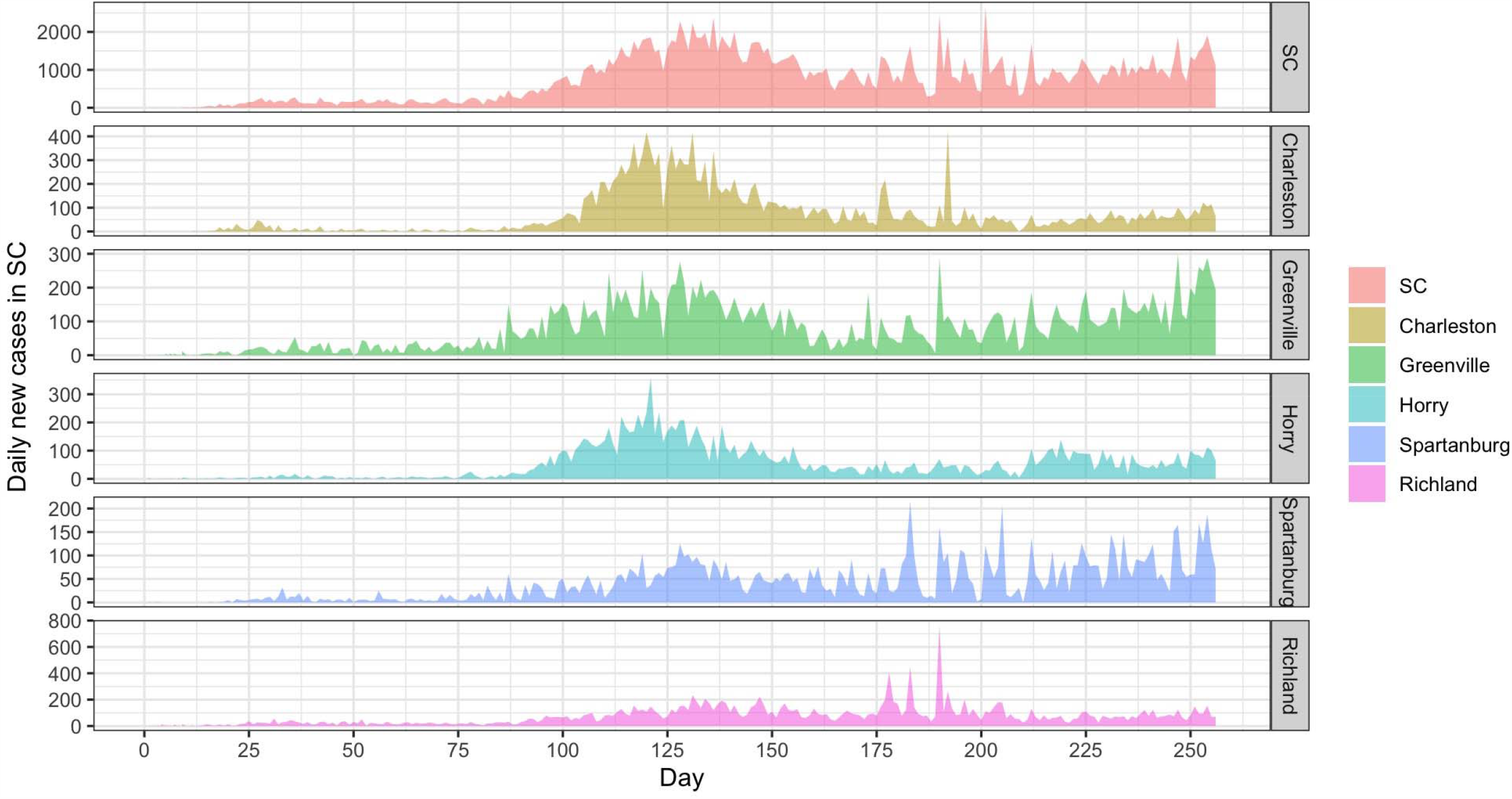
Daily COVID-19 new cases at both state- and county-levels in SC ***Note:*** SC: South Carolina.

At the county- level, the top five counties showed a similar trend of COVID-19 outbreaks and accounted for more than 40.0% of the total cases in SC. The daily new cases increased earlier in Greenville than the other four counties (i.e., Charleston, Horry, Spartanburg, and Richland).

Trends for population mobility at both state- and county- levels were similar. The numbers of people in SC (Twitter users in our data) with a moving distance of more than 0.5 mile decreased from 1,400 to 550 between March 6 and April 9, 2020. Although there were slight increases from the middle of April to that of June, the numbers were consistently around 1,000 after this timeframe. At the county-level, each of the five counties had less than 200 people with moving distance larger than 0.5 mile after the middle of March. Figure 2 shows the changes of population mobility at both state- and county- levels.

**Figure 2.**
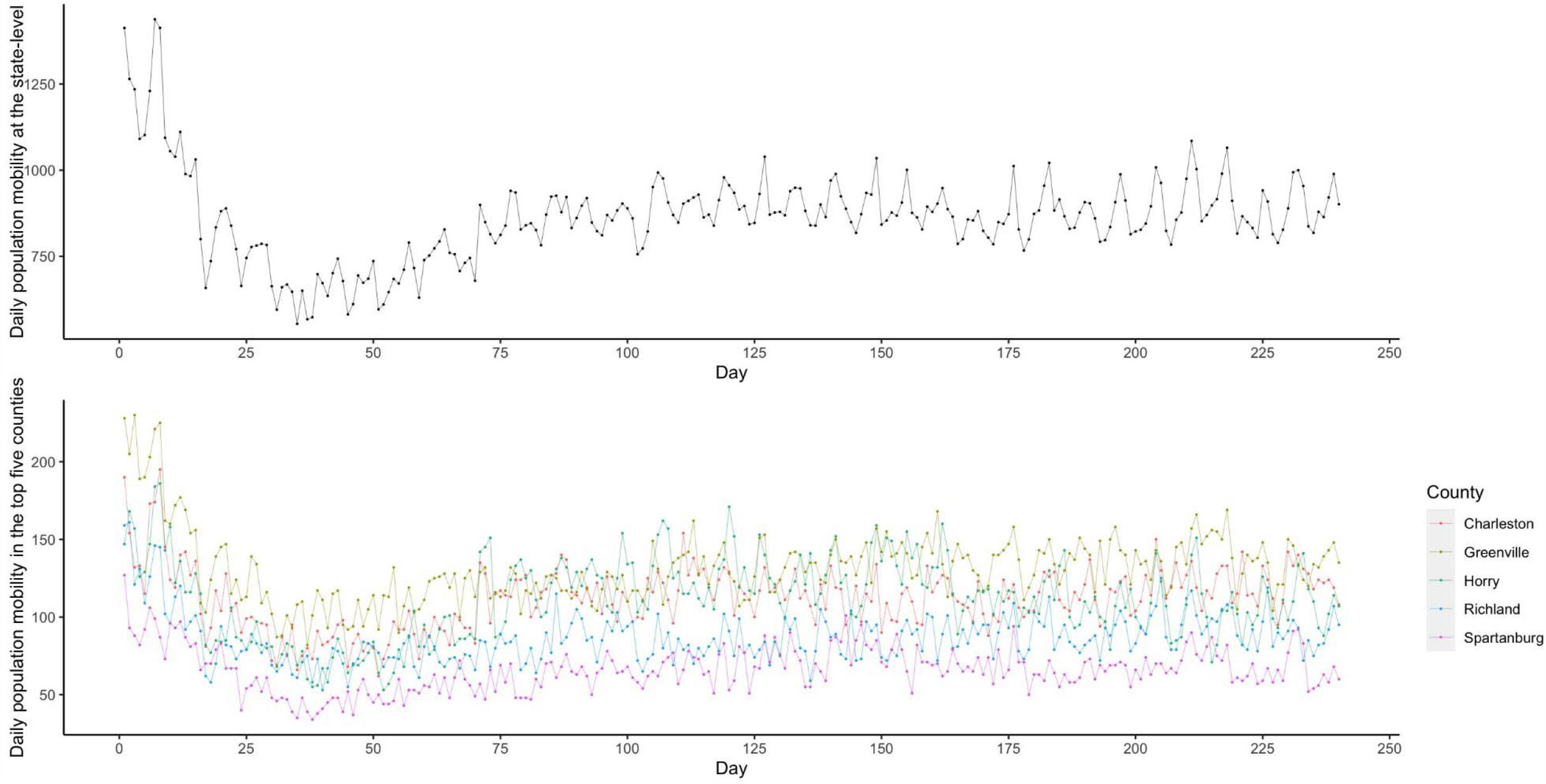
Daily population mobility at both state- and county-levels in SC ***Note:*** SC: South Carolina.

### Model selection of time series analyses

Following the model selection procedure, Poisson count time series model of COVID-19 incidence at the state-level was constructed using daily new cases and population mobility. Population mobility was positively associated with state-level COVID-19 daily new cases (β=0.818, 95%*CI*: 0.761∼0.876), and model using the past 7- day (1∼7 days) as time window had the smallest prediction error (Table 2). The prediction error of new cases in the next 3- day (235^th^∼ 237^th^) was 0.218.

**Table 2.**
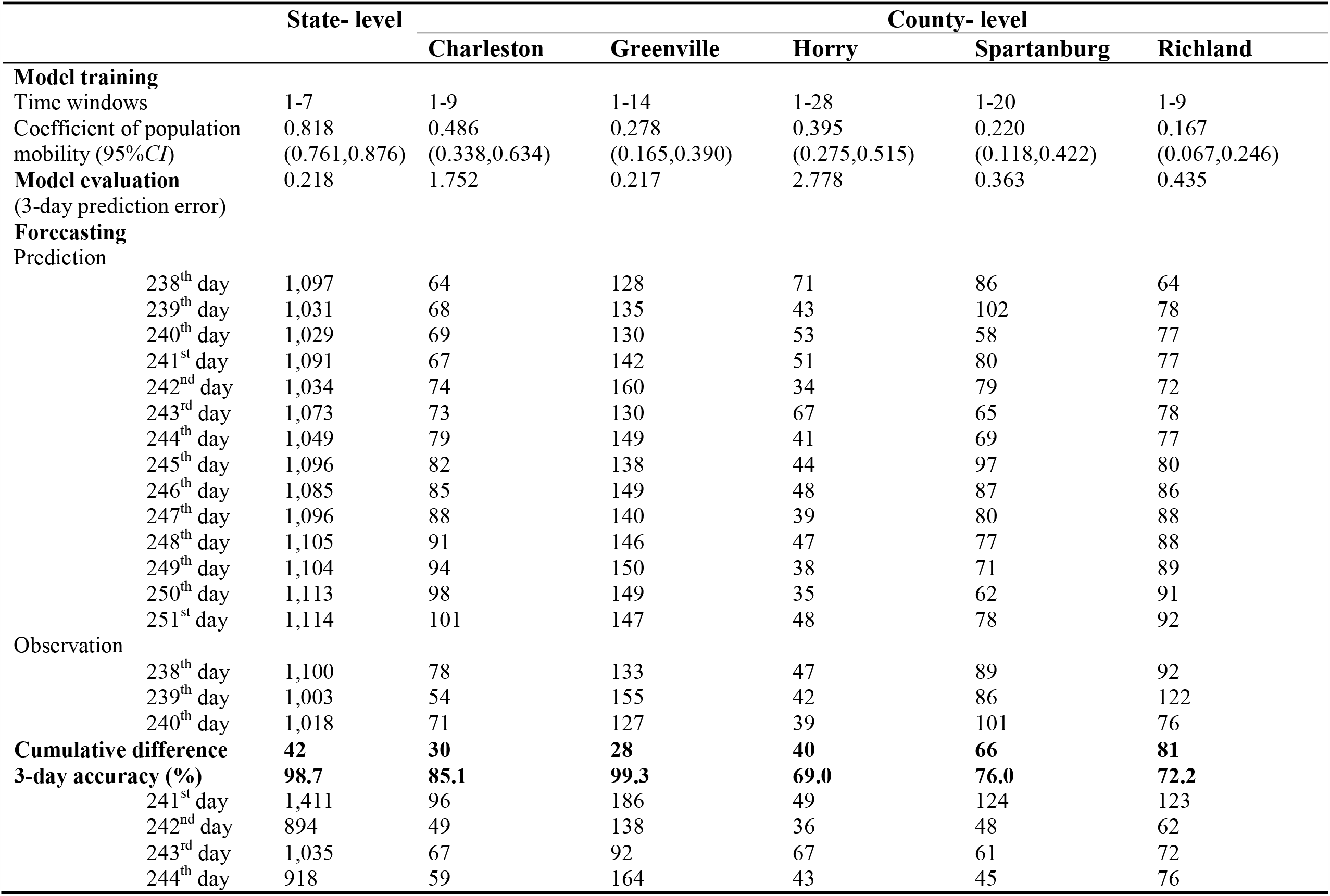
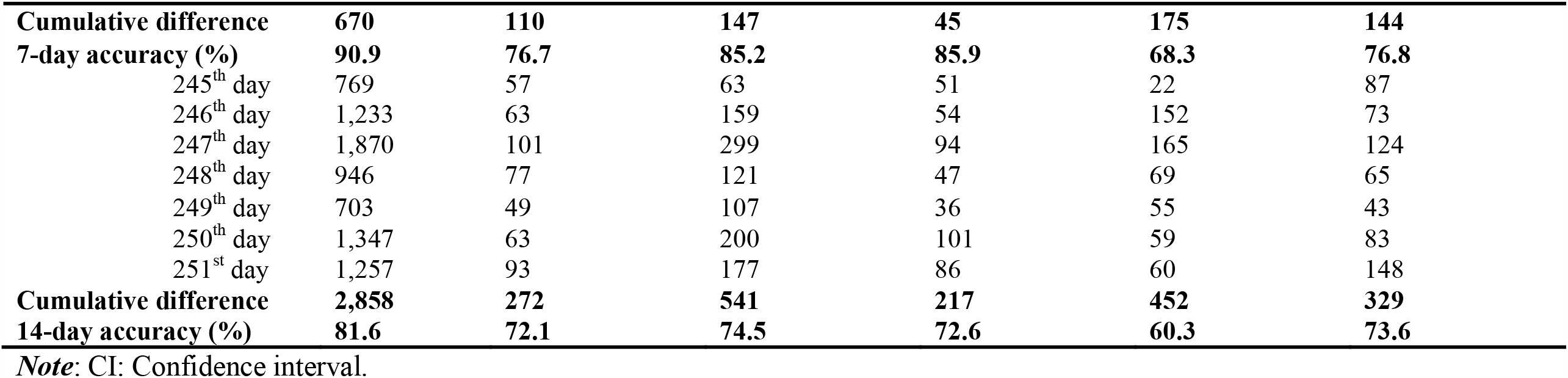
The impacts of population mobility on COVID-19 outbreaks in SC

At the county-level, a similar modelling procedure was employed. Population mobility was consistently and positively associated with COVID-19 new cases across the top five counties. The best time windows for Charleston, Greenville, Horry, Spartanburg, and Richland were 9-, 14-, 28-, 20-, and 9- days, respectively. Table 2 displays the detailed results of final model, correlation analysis, and 3-day prediction error at both state- and county- levels.

### COVID-19 daily new cases forecasting

Table 2 also presents the results of forecasting and prediction accuracy. Using final models with the selected time windows, COVID-19 daily new cases were forecasted for the next 14-day at both state- and county- levels. At the state- level, the 3-day cumulative difference and prediction accuracy were 42 and 98.7%, respectively. As compared to the 3-day predication accuracy, the 7- and 14- day accuracy reduced to 90.9% and 81.6%. At the county- level, among the top five counties, the 3-day prediction accuracy ranged from 69.0% to 99.3%. The prediction accuracy deceased in Charleston, Greenville, and Spartanburg with increased time span. In contrast, the prediction accuracy in Horry and Richland increased in 7-day prediction but decreased in 14-day prediction. The 14- day prediction accuracy among Horry and Richland were closer to their values in 3-day prediction.

## Discussion

This study leveraged disease surveillance data and Twitter-based population mobility to test the relationship between mobility and COVID-19 daily new cases and forecast the future transmission during the next 14 days at both state- and county- levels in SC. Results revealed that population mobility was significantly and positively associated with new daily COVID-19 cases. Using the selected models to forecast COVID-19 transmission, we found that although the prediction accuracy at state- level and most of the selected counties decreased as time span increased, the prediction accuracy remained acceptable. To the best of our knowledge, this is the first study that combined correlation analysis and forecasting together to investigate the impacts of population mobility on COVID-19 transmission at both state- and county- levels.

Population mobility could reflect the impacts of NPIs, reopening policies, and public holidays and estimate the social movement during the current COVID-19 pandemic. It is closely related to the COVID-19 outbreaks, which is in accordance with that of prior research [6,8,14- 16]. This study adds value to previous studies by examining the impacts of population mobility on COVID-19 incidence and evaluate its prediction efficacy at both state- and county- levels in SC during the two-week window. Although this indicator may only reflect the mobility among people who used Twitter, the results still revealed the positive a correlation between mobility and COVID-19 transmission.

Additionally, using Twitter-based mobility data to predict daily new COVID-19 cases could provide acceptable accuracy, which could also justify the validity and prediction efficacy of this indicator. The high prediction accuracy at the state-level was consistent with Wang’s finding in Arizona [16]. However, such a high prediction accuracy did not exist at the county-level. One possible explanation for this finding is that we did not capture or account for the influences of contextual factors (i.e., population density) and the roles of mitigating factors (e.g., wearing face mask, practicing social distancing) [16,20,21]. Additionally, the Twitter-based mobility did not differentiate the social movement at different locations, such as movement around parks, workplace, and retail places, which show different impacts on COVID-19 incidence [6]. Furthermore, in this study, we only captured population mobility at state- and county- levels while population mobility at zip code level might provide more accurate prediction. Finally, compared with mobility data from other platforms (e.g., Facebook, Google, Safegraph, Apple), our Twitter-based mobility indicator only estimated how many people with moving distance larger than a specific value. Nevertheless, the findings generated from this study confirmed the spatial-temporal relationship between Twitter-based mobility and COVID-19 outbreaks in SC as well as the prediction efficacy of population mobility.

Use of population mobility data has potential implications in future research and practices to curb COVID-19 outbreaks. From a research perspective, studies on mobility and COVID-19 could be studied at state-, county-, and/or zip code levels. In addition, mobility around different locations could provide detailed information regarding COVID-19 transmission, identify the most relevant mobility associated with daily new cases, and inform tailored interventions on social distancing by different locations to control disease outbreaks. Furthermore, the geospatial difference in the prediction accuracy of population mobility on daily new cases by county suggested that contextual factors, such as demographic characteristics and implementation fidelity of NPIs at county-level, should be accounted for in future research. Finally, since the incubation and transmission of COVID-19 are closely associated with time-varying factors, such as temperature and weather, such impacts should be accounted for in forecasting studies [22].

Regarding the practice of disease control and prevention, leveraging social media platform to monitor daily population mobility could improve the predictions of further COVID-19 transmission, inform proactive NPIs, and guide allocation of healthcare resources to reduce disease morbidity and mortality [23,24].

## Conclusions

Population mobility was positively associated with COVID-19 transmission at both state- and county- levels in SC. Using Twitter-based mobility data could provide acceptable prediction for COVID-19 daily new cases. The application of social media platforms to monitor population mobility and predict COVID-19 spread could inform proactive measures to curb disease outbreaks and plan coordinated responses.

## Data Availability

The authors agree to share the data upon request.

https://github.com/nytimes/covid-19-data

## Acknowledgements

This study was supported by the National Institute of Health (NIH) Research Grant R01AI127203-01A by National Institute of Allergy and Infectious Diseases and National Science Foundation (NSF) Grant No. 2028791.

## Conflicts of interest

None declared.

## Abbreviations

COVID-19: Coronavirus disease 2019
NIH: National Institute of Health
NPIs: Non-pharmaceutical interventions
NSF: National Science Foundation
SC: South Carolina
US: United States

## Notes

### Competing Interest Statement

The authors have declared no competing interest.

### Author Declarations

The study protocol was approved by the Institutional Review Boards at the University of South Carolina.

